# Clonal Hematopoiesis of Indeterminate Potential is Associated with Acute Kidney Injury

**DOI:** 10.1101/2023.05.16.23290051

**Authors:** Caitlyn Vlasschaert, Cassianne Robinson-Cohen, Bryan Kestenbaum, Samuel A. Silver, Jian-Chun Chen, Elvis Akwo, Pavan K Bhatraju, Ming-Zhi Zhang, Shirong Cao, Ming Jiang, Yinqiu Wang, Aolei Niu, Edward Siew, Holly J Kramer, Anna Kottgen, Nora Franceschini, Bruce M. Psaty, Russell P. Tracy, Alvaro Alonso, Dan E. Arking, Josef Coresh, Christie M Ballantyne, Eric Boerwinkle, Morgan Grams, Matthew B. Lanktree, Michael J. Rauh, Raymond C. Harris, Alexander G. Bick

**Affiliations:** Department of Medicine, Queen’s University, Kingston, Ontario, Canada; Division of Nephrology and Hypertension, Department of Medicine, Vanderbilt O’Brien Center for Kidney Disease, School of Medicine, Vanderbilt University, Nashville, Tennessee; Kidney Research Institute, Division of Nephrology, Department of Medicine, University of Washington, Seattle, Washington; Division of Pulmonary, Critical Care and Sleep Medicine, Department of Medicine, University of Washington, Seattle, Washington; Departments of Public Health Sciences and Medicine, Loyola University Chicago, Maywood, Illinois, USA; Institute of Genetic Epidemiology, Faculty of Medicine and Medical Center, University of Freiburg, Freiburg, Germany; Department of Epidemiology, Bloomberg School of Public Health, Johns Hopkins University, Baltimore, Maryland; Department of Epidemiology, Gillings School of Global Public Health, University of North Carolina, Chapel Hill, North Carolina; Cardiovascular Health Research Unit, Departments of Medicine, Epidemiology and Health Systems and Population Health, University of Washington, Seattle, WA, USA; Kaiser Permanente Washington Health Research Institute, Seattle, WA, USA; Pathology and Biochemistry, University of Vermont, Burlington, Vermont, USA; Department of Epidemiology, Rollins School of Public Health, Emory University, Atlanta, Georgia; McKusick-Nathans Institute, Department of Genetic Medicine, John Hopkins University School of Medicine, Baltimore, MD; Welch Center for Prevention, Epidemiology, and Clinical Research, Johns Hopkins University, Baltimore, MD; Department of Medicine, Baylor College of Medicine, Houston, TX, USA; Human Genetics Center, The University of Texas Health Science Center at Houston, Houston, TX 77030, USA; Division of Nephrology, Department of Internal Medicine, Johns Hopkins University, Baltimore, MD; Department of Medicine and Department of Health Research Methods, Evidence and Impact, McMaster University, Hamilton, Ontario, Canada; St. Joseph’s Healthcare Hamilton, Hamilton, Ontario, Canada; Population Health Research Institute, Hamilton, Ontario, Canada; Department of Pathology and Molecular Medicine, Queen’s University, Kingston, Ontario, Canada; Department of Veterans Affairs, Nashville, Tennessee; Division of Genetic Medicine, Department of Medicine, School of Medicine, Vanderbilt University, Nashville, Tennessee

**Author notes:** Correspondence to Dr. Bick and Dr. Harris.

## Abstract

Age is a predominant risk factor for acute kidney injury (AKI), yet the biological mechanisms underlying this risk are largely unknown and to date no genetic mechanisms for AKI have been established. Clonal hematopoiesis of indeterminate potential (CHIP) is a recently recognized biological mechanism conferring risk of several chronic aging diseases including cardiovascular disease, pulmonary disease and liver disease. In CHIP, blood stem cells acquire mutations in myeloid cancer driver genes such as *DNMT3A, TET2, ASXL1* and *JAK2* and the myeloid progeny of these mutated cells contribute to end-organ damage through inflammatory dysregulation. We sought to establish whether CHIP causes acute kidney injury (AKI). To address this question, we first evaluated associations with incident AKI events in three population-based epidemiology cohorts (N = 442,153). We found that CHIP was associated with a greater risk of AKI (adjusted HR 1.26, 95% CI: 1.19–1.34, p<0.0001), which was more pronounced in patients with AKI requiring dialysis (adjusted HR 1.65, 95% CI: 1.24–2.20, p=0.001). The risk was particularly high in the subset of individuals where CHIP was driven by mutations in genes other than *DNMT3A* (HR: 1.49, 95% CI: 1.37–1.61, p<0.0001). We then examined the association between CHIP and recovery from AKI in the ASSESS-AKI cohort and identified that non-*DNMT3A* CHIP was more common among those with a non-resolving pattern of injury (HR 2.3, 95% CI: 1.14–4.64, p = 0.03). To gain mechanistic insight, we evaluated the role of *Tet2*-CHIP to AKI in ischemia-reperfusion injury (IRI) and unilateral ureteral obstruction (UUO) mouse models. In both models, we observed more severe AKI and greater post-AKI kidney fibrosis in *Tet2*-CHIP mice. Kidney macrophage infiltration was markedly increased in *Tet2*-CHIP mice and *Tet2*-CHIP mutant renal macrophages displayed greater pro-inflammatory responses. In summary, this work establishes CHIP as a genetic mechanism conferring risk of AKI and impaired kidney function recovery following AKI via an aberrant inflammatory response in CHIP derived renal macrophages.

## Introduction

Acute kidney injury (AKI) affects more than 1 in 5 hospitalized adults worldwide^1,2^, and is associated with significant health care costs and patient mortality, greater than that of heart failure or diabetes.^3^ AKI is characterized by an inflammatory and fibrotic response to an initial insult, most commonly kidney hypoperfusion, leading to a quantifiable impairment in kidney function based on serum markers and urine output.^4^ Following AKI, there is a remarkable heterogeneity of outcomes. Only about half of AKI cases return to baseline kidney function within 3 months, while many have residual kidney damage.^5^ Recognized patient factors that predispose to AKI and to progression from AKI to chronic kidney disease largely consist non-modifiable clinical risk factors such as age.^2^ To date, there have been no identified genetic factors that predispose to AKI or AKI outcomes.

Dysregulated inflammatory responses in macrophages and other inflammatory cells can occur in the setting of clonal hematopoiesis of indeterminate potential (CHIP), a common age-related hematologic process characterized by the clonal expansion of hematopoietic stem cells (HSCs) and their progeny following an acquired genetic mutation (commonly in *DNMT3A, TET2, ASXL1* and *JAK2*). While less than 0.5% of CHIP cases per year progress to overt hematologic cancer^6,7^, CHIP is associated with an estimated 40% greater risk of mortality^8^ largely due to disease beyond the hematopoietic system including cardiovascular disease^9–12^, pulmonary disease^13,14^, liver disease^15^, and other inflammatory conditions.^16–19^ CHIP may influence kidney health as experimental recapitulation of CHIP by transplanting a small fraction of HSCs with pathogenic *Tet2* mutations in mice have shown that the *Tet2*-deficient cells readily replace resident macrophage populations in the kidney and liver^15,20^, and myeloid cells play pivotal roles in response to injury, repair and management of the kidney microenvironment.^21–23^

Here, we tested the hypothesis that CHIP is a risk factor for AKI. We first show that CHIP is associated with incident AKI in three large population-based cohorts and is more pronounced for non-*DNMT3A* CHIP. We then show that non-*DNMT3A* CHIP is associated with a non-resolving AKI pattern in the ASSESS-AKI cohort. Finally, we show that AKI severity is more pronounced and AKI recovery is impaired in a mouse model with *Tet2*-CHIP in both the ischemia-reperfusion injury (IRI) and unilateral ureteral obstruction (UUO) AKI models, with risk mediated by an aberrant inflammatory response in *Tet2*-CHIP derived renal macrophages.

## Results

### CHIP and incident AKI in the UK Biobank

We first assessed the association between CHIP and incident AKI in the UKB. The mean baseline age was 57 ± 8 (SD) years, the mean baseline eGFR was 95 ± 14 ml/min/1.73m^2^ (**Table 1**). The prevalence of CHIP was 3.4% and increased with age (**Extended Data Figure 1**). *DNMT3A* was the most commonly mutated gene followed by *TET2* and *ASXL1*. There were 15,736 incident AKI events among 428,793 participants (3.1 events per 1000 person-years).

**Table 1.** Baseline cohort characteristics for incident AKI studies.

|  | **UK Biobank**<br>(N = 428,793) |  | **ARIC**<br>(N = 10,570) |  | **CHS**<br>(N = 2,790) |  |
| --- | --- | --- | --- | --- | --- | --- |
|  | *CHIP* | *No CHIP* | *CHIP* | *No CHIP* | *CHIP* | *No CHIP* |
| Participants (N) | 14,552 | 414,241 | 743 | 9,827 | 404 | 2,386 |
| Age (years; mean ± SD) | 61 ± 7 | 56 ± 8 | 60 ± 6 | 58 ± 6 | 75 ± 6 | 74 ± 6 |
| Male sex (%) | 47 | 46 | 48 | 45 | 47 | 43 |
| eGFR (ml/min/1.73m<sup>2</sup>; mean ± SD) | 91 ± 15 | 95 ± 14 | 93 ± 16 | 96 ± 15 | 66 ± 16 | 69 ± 16 |
| Diabetes (%) | 2 | 1 | 10 | 10 | 19 | 16 |
| Hypertension (%) | 8 | 6 | 4 | 4 | 47 | 46 |
| Smoking (%) | 54 | 41 | 62 | 57 | 54 | 55 |

CHIP was associated with a 34% greater risk of incident AKI (HR 1.34, 95% confidence interval (CI): 1.29– 1.40, p<0.0001) in a Cox proportional hazards model in fully adjusted analyses (**Figure 1a**). The association between CHIP and AKI was stronger when AKI was limited to cases receiving dialysis (AKI-D; HR 1.65, 95% CI: 1.24 – 2.20, p=0.001). The risk for AKI associated with CHIP was also higher in individuals with mutations in genes other than *DNMT3A* (**Figure 1b**; HR 1.54, 95% CI: 1.41–1.68 for any AKI and HR 2.18, 95% CI: 1.51–3.15 for AKI-D, p<0.0001). Although the absolute risk for AKI was higher among those with baseline CKD, CHIP conferred a similar absolute risk difference among those with and those without baseline CKD (**Extended Data Figure 2**).

**Figure 1.**
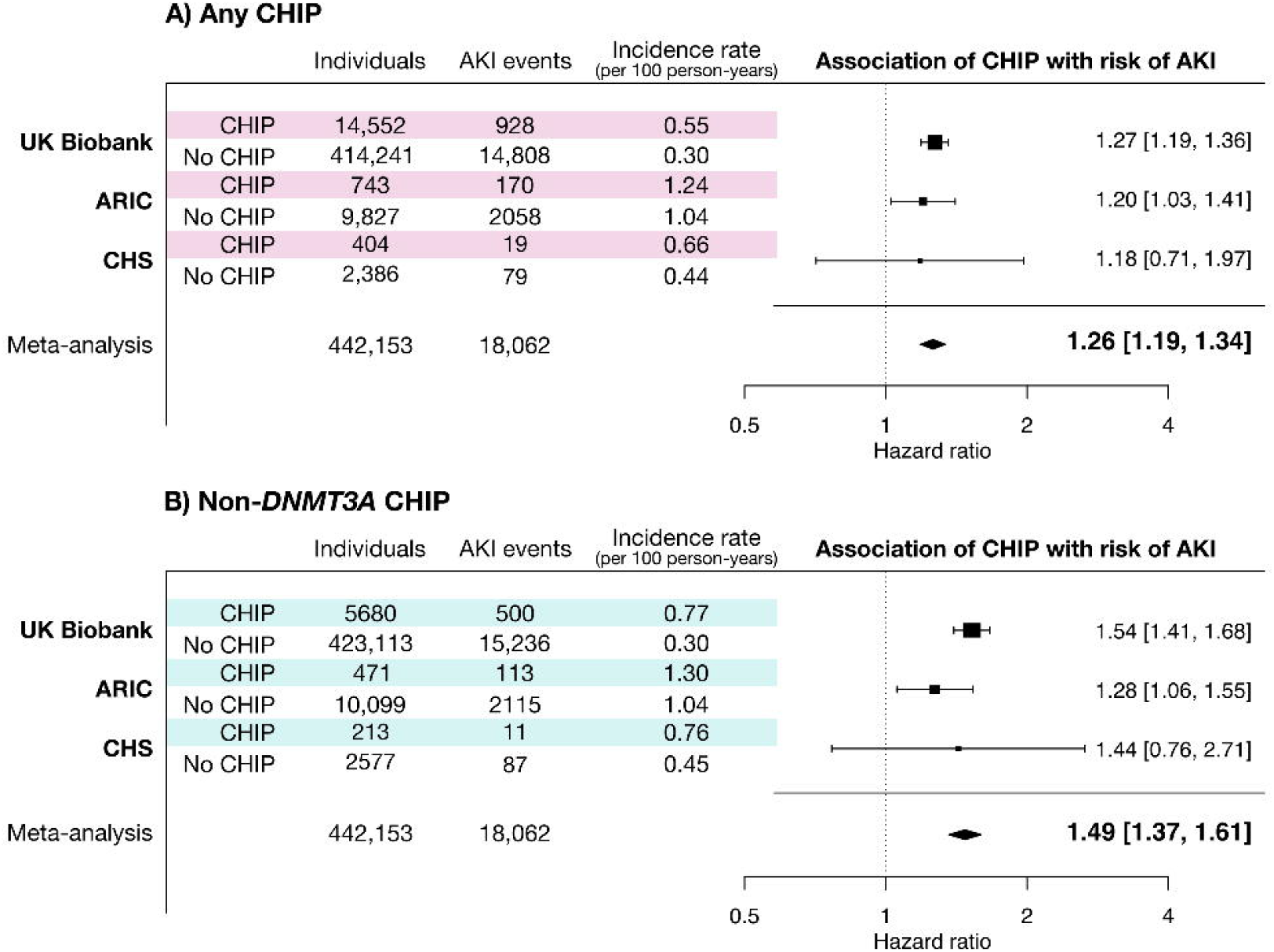
CHIP is associated with a greater risk of incident AKI in three population-based cohorts. **A)** Incident AKI risk was assessed in the UK Biobank, where events were abstracted from ICD codes during hospital discharges, and in two smaller population-based cohorts where additional manual curation was performed to ascertain AKI events. **B)** The risk of AKI is greater for CHIP driven by mutations in genes other than *DNMT3A* (non-*DNMT3A* CHIP). All analyses were adjusted for age, age^2^, sex, baseline eGFR, baseline smoking status, diabetes, and hypertension, as well as either 10 PCs of genetic ancestry (UKB) or self-reported race/ethnicity (ARIC & CHS).

### CHIP and incident AKI in Atherosclerosis Risk in Community and Cardiovascular Health Study cohorts

Next, we assessed the association of CHIP with incident AKI in two prospective cohort studies: the Atherosclerosis Risk in Community (ARIC) cohort and the Cardiovascular Health Study (CHS).^25,26^ The mean baseline age was 57 ± 4 (SD) years in the ARIC cohort and 72 ± 5 years in the CHS cohort. Mean baseline eGFR was 96 ± 15 ml/min/1.73m^2^ in ARIC and 68 ± 16 ml/min/1.73m^2^ in CHS. As previously reported, the prevalence of CHIP was 7.6% in ARIC and 14.5% in CHS^27^ (age distribution shown in **Extended Data Figure 1**). The baseline characteristics for these cohorts are listed in Table 1.

There were 2,228 events among 10,570 individuals in ARIC (10.5 events per 1000 p-y), and 98 events among 2,790 individuals in CHS (4.7 events per 1000 p-y). CHIP was associated with a 20% greater risk of AKI in a meta-analysis of these cohorts (HR 1.20; 95% CI: 1.03–1.39, **Figure 1a**), and, consistent with what was observed in the UKB, the point estimate of the magnitude of relative risk for AKI was higher for individuals with non-*DNMT3A* CHIP (**Figure 1b**; meta-analyzed HR 1.29, 95% CI: 1.08–1.55).

### CHIP and recovery from AKI in the ASSESS-AKI cohort

We assessed whether CHIP was associated with patterns of AKI recovery in the ASSESS-AKI cohort, which enrolled 769 individuals with AKI events during a hospitalization and longitudinally tracked their clinical outcomes over 5 years.^28^ Among 321 with AKI, 74 (23%) had CHIP. Among the individuals with AKI, non-*DNMT3A* CHIP and large CHIP clones (VAF ≥ 10%) were more than twice as common among individuals with a non-resolving AKI pattern compared to those with resolving AKI (**Figure 2b**), including after adjusting for age and other relevant covariates (**Figure 2c**). Additionally, large CHIP clones were associated with an increased risk of the study primary outcome (incident kidney failure or 50% decline in eGFR over 5 years; HR 2.9, 95% CI: 1.1-8.0; **Figure 2d**).

**Figure 2.**
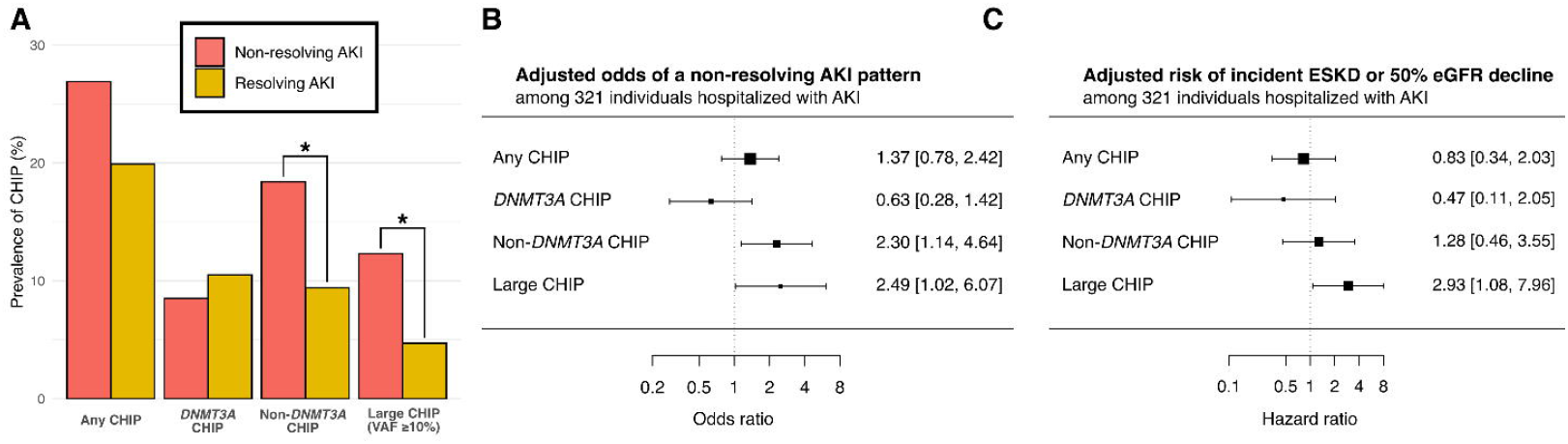
CHIP is associated with impaired recovery from AKI in the ASSESS-AKI study. **A)** Prevalence of CHIP among individuals with a resolving AKI pattern (n = 191) compared to a non-resolving AKI pattern (n = 130), as defined by Bhatraju *et al*.^5^ **B)** Odds of non-resolving AKI pattern by CHIP status, adjusted for age, sex, baseline creatinine, AKI stage, smoking status, race, and history of diabetes, hypertension, and cardiovascular disease. **C)** Risk of significant kidney function impairment (primary study composite outcome of ESKD or eGFR decline by ≥50%) over 5 years of follow-up among ASSESS-AKI participants with baseline AKI by CHIP status, adjusted for age, sex, baseline creatinine, AKI stage, smoking status, race, and history of diabetes, hypertension, and cardiovascular disease.

### CHIP and AKI severity in mouse models

We sought to leverage CHIP mouse models to obtain mechanistic insights into how CHIP contributed to AKI severity. Since non-*DNMT3A*-CHIP was most strongly associated with AKI outcomes in our epidemiologic studies, we generated a mouse model of *TET2*-CHIP, the most common type of non-*DNMT3A* CHIP. Briefly, we performed a bone marrow transplant containing 20% CD45.2^+^ *Tet2*^-/-^ cells and 80% CD45.1^+^ *Tet2*^+/+^ cells in lethally irradiated mice (**Extended Data Figure 3a**). Control mice received a bone marrow transplant (BMT) of 20% CD45.2^+^ *Tet2*^+/+^ cells and 80% CD45.1^+^ *Tet2*^+/+^ cells. These mice are referred to as *Tet2*^*-/-*^ and wild type (WT), respectively, throughout the text. When studied 10 weeks after bone marrow transplantation, mice receiving the *Tet2* WT CD45.2 cells had a minority of both kidney macrophage and neutrophil CD45.2 cells in control kidneys (**Extended Data Figure 3c & d**). In contrast, the *Tet2*^*-/-*^ mice had a significant increase of *Tet2*^*-/-*^ cells in the intrinsic myeloid kidney cell population.

*Tet2*^*-/-*^ mice had an exaggerated AKI pattern following ischemia reperfusion injury (IRI). BUN increased within 24 hours to approximately 75 mg/dl in WT mice and decreased over the subsequent 8 days, whereas the BUN increases following the same ischemic insult in *Tet2*^*-/-*^ mice were significantly higher (p<0.01, **Figure 3a**).

**Figure 3.**
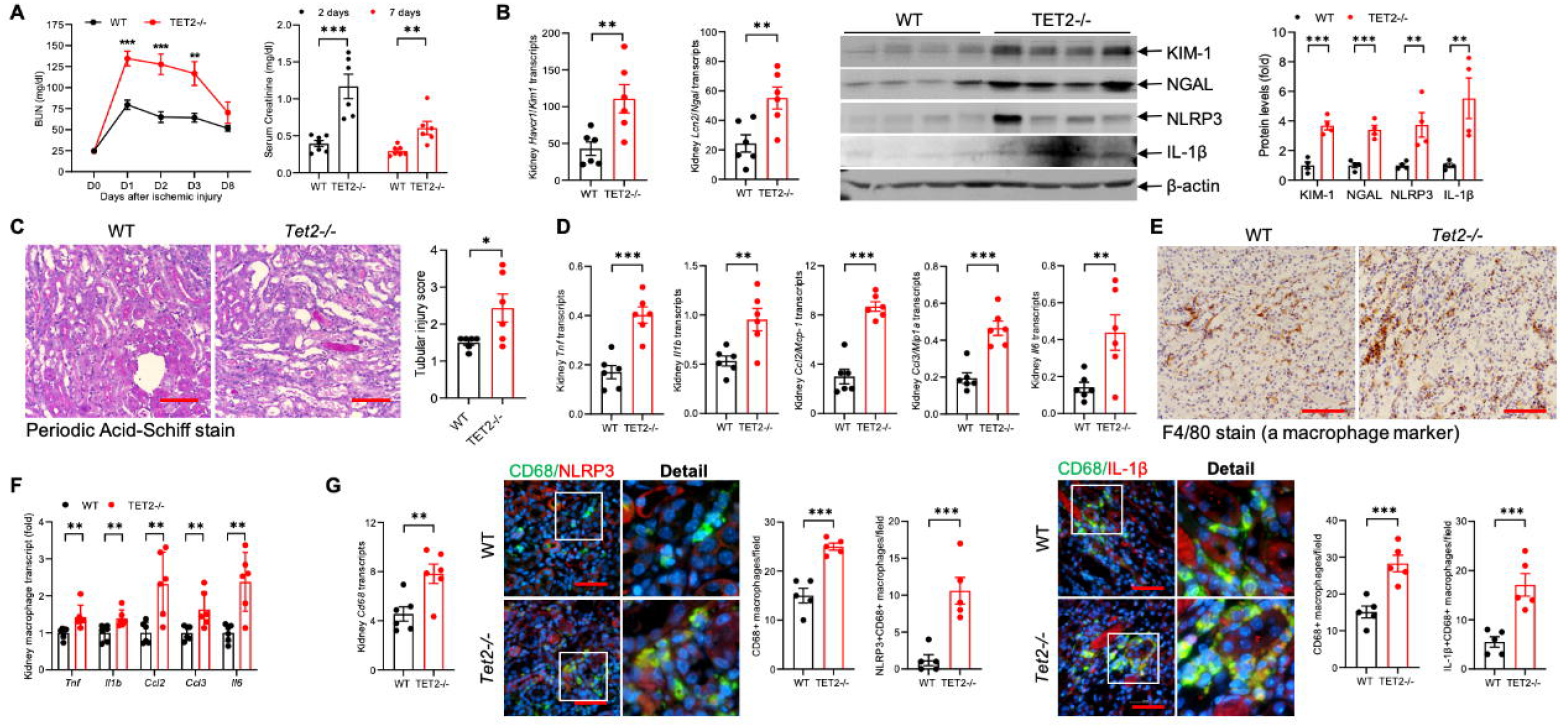
Early response of hematopoietic deletion of *Tet2*^*-/-*^ to ischemic kidney injury. Mice were subjected to 32 minutes of ischemic injury to the left kidney with simultaneous removal of the right kidney. **A**) BUN levels and serum creatinine were significantly higher in *Tet2*^*-/-*^ mice. **B**) In kidneys of *Tet2*^*-/-*^ mice 8 days after injury, there was increased mRNA and immunoreactive protein kidney levels of the tubule injury markers Kim-1 and Ngal and inflammatory markers, NLRP3 and IL-1β. **C**) The tubule injury score was significantly higher in *Tet2*^*-/-*^ mice. **D**) *Tet2*^*-/-*^ kidneys had increased mRNA expression of proinflammatory cytokines, **E**) increased macrophage infiltration, indicated by increased immunoreactive expression of the macrophage marker F4/80, and **F**) isolated macrophages had increased mRNA expression of proinflammatory macrophages. **G**) There was increased kidney expression of *Cd68* mRNA and increased CD68 colocalization with immunoreactive NLRP3 and IL-β. *p<0.05; **p<0.01; scale bar=50 µm.

Similarly, serum creatinine was also significantly higher in *Tet2*^*-/-*^ mice than WT mice at both 2 and 7 days after ischemic injury. When mice were subjected to more extensive ischemic injury, there was increased early mortality in *Tet2*^*-/-*^ mice (4 of 6) compared to WT mice (1 of 7; **Extended Data Figure 4**) consistent with increased sensitivity to ischemic injury.

*Tet2*^*-/-*^ mice had an exaggerated histologic and molecular AKI pattern following IRI. 8 days after ischemic injury, kidneys of *Tet2*^*-/-*^ mice had increased expression of mRNA and protein for the tubule injury markers, KIM-1 and NGAL (**Figure 3b**), and histologic analysis indicated significantly more tubule injury (**Figure 3c**). Kidneys of *Tet2*^*-/-*^ mice had increased mRNA for proinflammatory cytokines *Tnf, Il6, Il1b*, and chemokines *Ccl2* and *Ccl3* (**Figure 3d**) as well as increased immunostaining for the macrophage marker F4/80 (**Figure 3e**).

*Tet2*^*-/-*^ pathology was localized to the *Tet2*^*-/-*^ mutant renal macrophages. Macrophages isolated from the kidneys of *Tet2*^*-/-*^ mice had increased mRNA expression of proinflammatory cytokines *Tnf, Il6, Il1b* both at baseline (**Extended Data Figure 3e**) and after ischemic injury (**Figure 3f**). Compared to WT mice, *Tet2*^*-/-*^ mice also had increased kidney mRNA and immunoreactive expression of the macrophage inflammatory marker, CD68 (**Figure 3g**) and expressed higher levels of the NLRP3 inflammasome and its product IL-1β (**Figure 3b**), both of which colocalized with CD68-positive macrophages (**Figure 3g**). Importantly, the increased co-expression was only seen in the CD45.2 (*Tet2*^*-/-*^) cells of *Tet2*^*-/-*^ mice and not in the CD45.1 (*Tet2*^*+/+*^) cells of *Tet2*^*-/-*^ mice, nor in either cell type in the WT mice (**Extended Data Figure 5**). At this timepoint, the kidneys of *Tet2*^*-/-*^ mice also had increased mRNA levels of profibrotic (*Tgfb, Ctgf, Acta2*) and extracellular matrix-associated *(Col1a1, Col3a1, Fn and Vim)* genes compared to WT mice (**Figure 4a**).

**Figure 4.**
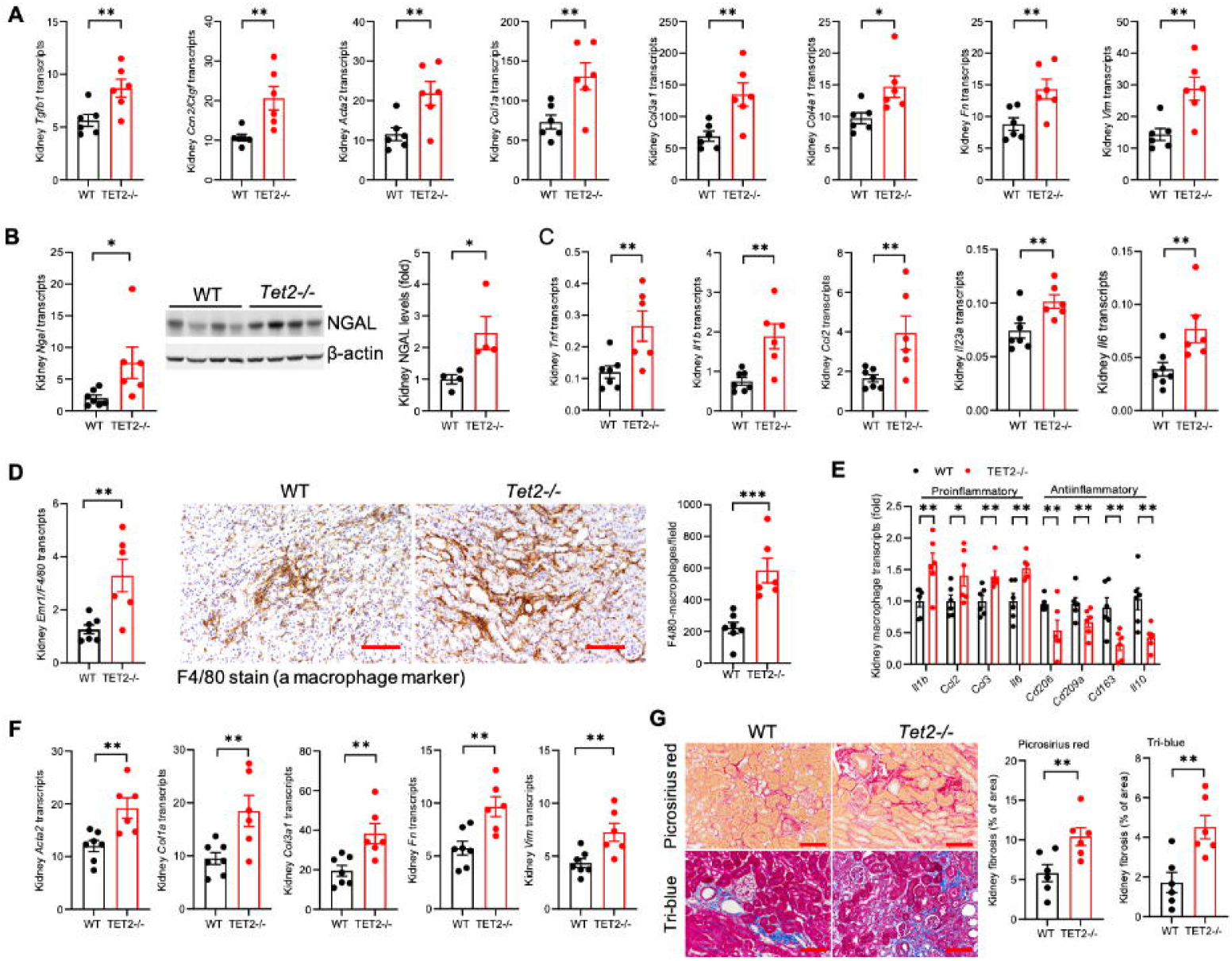
Increased kidney interstitial fibrosis following ischemic kidney injury with hematopoietic deletion of *Tet2*. **A)** 8 days after injury, kidneys of *Tet2*^*-/-*^ mice expressed increased profibrotic markers. **B**) 28 days after injury, kidneys of *Tet2*^*-/-*^ mice still expressed increased KIM-1 and NGAL and **C**) increased mRNA for inflammatory cytokines. **D**) *Tet2*^*-/-*^ kidneys had increased macrophage infiltration, and **E**) kidney macrophages isolated from *Tet2*^*-/-*^ kidneys expressed increased mRNA for increased proinflammatory and decreased mRNA for anti-inflammatory, pro-reparative cytokines. **F**) 28 days after injury, *Tet2*^*-/-*^ kidneys expressed increased mRNA for fibrotic markers and **G**) had increased Picrosirius red and Masson blue staining for interstitial collagens. *p<0.05; **p<0.01; scale bar=50 µm

*Tet2*^*-/-*^ mice recapitulated non-resolving AKI pathology. Twenty-eight days after ischemic injury, kidneys from *Tet2*^*-/-*^ mice maintained elevated mRNA and protein levels of kidney injury markers (**Figure 4b**), increased mRNA for proinflammatory cytokines (**Figure 4c**), and ongoing infiltration of leukocytes, as demonstrated by increased F4/80 mRNA and immunostaining for macrophages (**Figure 4d**), increased mRNA for *Ly6g* (neutrophils), and *cd4* and *cd8* T-cells; (**Extended Data Figure 6**). Isolated kidney macrophages expressed increased mRNA for proinflammatory genes and decreased mRNA of pro-recovery (“M2”) genes (**Figure 4e**). Kidneys from *Tet2*^*-/-*^ mice also maintained elevated mRNA expression of extracellular matrix genes (**Figure 4f**) and developed significantly increased interstitial fibrosis, determined by Picrosirius red and Masson trichrome staining (**Figure 4g**).

We sought to recapitulate the AKI phenotype in an independent mouse model, unilateral ureteral obstruction. Comparing WT and *Tet2*^*-/-*^ mice at 7 days, the affected kidney of *Tet2*^*-/-*^ mice had increased mRNA expression of proinflammatory cytokines (*Il1b, Tnf, ccl3, ccl2*) (**Extended Data Figure 7A**), increased kidney macrophage (F4/80) and neutrophil (Ly6G) invasion (**Extended Data Figure 7B**), and significantly increased interstitial fibrosis (**Extended Data Figure 7C**).

## Discussion

Here by integrating large scale human genetic data from four human epidemiologic studies and two distinct *in vivo* models, we identify that CHIP is associated with AKI and poor kidney outcomes following AKI via an aberrant renal macrophage inflammatory response. The nearly two-fold increased risk of AKI progressing to dialysis and non-resolving AKI we observe in non-*DNMT3A* CHIP clones is similar in magnitude to that observed for CHIP and cardiovascular disease. As this association was independent of age, sex, smoking status, baseline eGFR and presence of diabetes and hypertension, with effect estimates were remarkably consistent across cohorts, despite notable differences in baseline characteristics and AKI incidence, we suggest that this is the first genetic risk factor that has been associated with AKI.

The association between CHIP and incident AKI events may reflect increased predisposition to AKI, greater severity of AKI, and/or impaired recovery from AKI. In order to differentiate these mechanisms, we assessed short- and long-term post-AKI outcomes by CHIP status in participants enrolled in the ASSESS-AKI study. Though limited by small sample size, we showed that non-*DNMT3A* CHIP was associated with a non-resolving AKI pattern. Additionally, since ASSESS-AKI participant CHIP status was ascertained using a targeted sequencing method that is highly sensitive to small CHIP clones, we conducted a sensitivity analysis examining only large clones with VAF ≥ 10%. We identified that large CHIP clones were associated both with a non-resolving AKI pattern and with higher rates of kidney failure over 5 years of study follow-up. These data suggest that non-*DNMT3A* CHIP is associated with impaired recovery from AKI in human subjects.

To demonstrate that non-*DNMT3A* CHIP is causally associated with AKI and further delineate disease mechanisms, we focused on inactivating mutations in *TET2* CHIP, which makes up approximately 30% of the non-*DNMT3A* CHIP cases in our study. After experimental ischemic or obstructive kidney injury, the *Tet2-*CHIP mice displayed more severe AKI and impaired post-AKI recovery when compared to mice with intact *Tet2*, as evidenced by higher serum levels of markers of impaired kidney function and injury (Cr and BUN; KIM-1 and NGAL) as well as structural kidney damage including higher more pronounced initial tubular injury and kidney interstitial fibrosis at 28-days post-AKI.

Greater local inflammation by infiltrating pro-inflammatory macrophages appears to underlie the worse post-AKI outcomes in *Tet2*-CHIP mice. We observed elevated levels of several inflammatory cytokines and chemokines in the kidneys of *Tet2*-CHIP mice, which primarily derived from kidney-infiltrating macrophages. Additionally, we showed that infiltrating *Tet2*^*-/-*^ renal macrophages but not *Tet2*^*+/+*^ renal macrophages had heightened NLRP3 inflammasome activation and IL-1β production. We and others have previously observed that *Tet2*^*-/-*^ derived macrophages are similarly hyperactivated in other tissue contexts.^9,10,15,17,18,29^ Overall, our study suggests that CHIP plays an inhibitory role in recovery after AKI due to increased pro-inflammatory signaling by mutated infiltrating macrophages.

In conclusion, CHIP is associated with an elevated risk of AKI specifically through the promotion of renal macrophage inflammation. Through targeting of the NLRP3 inflammasome or downstream mediators, CHIP may be a modifiable risk factor for AKI and progression to end stage kidney disease.

## Data Availability

All data produced are available online to registered UK Biobank users or upon reasonable request to the authors.

## Acknowledgements

This work was funded by a National Institute of Diabetes and Digestive and Kidney Diseases (NIDDK) grant #R01DK132155 awarded to C.R.C., R.J.H, and A.G.B. Support for whole-genome sequencing for the TOPMed program provided by the National Heart, Lung, and Blood Institute (NHLBI). Genome sequencing for “NHLBI TOPMed: the Atherosclerosis Risk in Communities Study” (phs001211.v1.p1) was performed at the Broad Institute Genomics Platform (3U54HG003273-12S2, HHSN268201500015C, 3R01HL092577-06S1). Genome sequencing for “NHLBI TOPMed: the Cardiovascular Health Study” (phs001368.v1.p1) was performed at the Broad Institute Genomics Platform (HHSN268201600034I) and Baylor College of Medicine Human Genome Sequencing Center (HHSN268201600033I). Centralized read mapping and genotype calling along with variant quality metrics and filtering were provided by the TOPMed Informatics Research Center (3R01HL-117626-02S1; contract HHSN268201800002I). Phenotype harmonization, data management, sample-identity QC, and general study coordination were provided by the TOPMed Data Coordinating Center (3R01HL-120393-02S1; contract HHSN268201800001I). This research was supported by contracts HHSN268201200036C, HHSN268200800007C, HHSN268201800001C, N01HC55222, N01HC85079, N01HC85080, N01HC85081, N01HC85082, N01HC85083, N01HC85086, 75N92021D00006, and grants U01HL080295 and U01HL130114 from the National Heart, Lung, and Blood Institute (NHLBI), with additional contribution from the National Institute of Neurological Disorders and Stroke (NINDS). Additional support was provided by R01AG023629 from the National Institute on Aging (NIA) and NHLBI grant R01148050. A full list of principal ARIC and CHS investigators and institutions can be found at https://sites.cscc.unc.edu/aric/ and CHS-NHLBI.org, respectively. The content is solely the responsibility of the authors and does not necessarily represent the official views of the National Institutes of Health. The funders did not have a role in study design, data collection, analysis, reporting, or the decision to submit for publication.

## Methods

### CHIP ascertainment

Acquired DNA mutations meeting established criteria for CHIP were identified as previously described in the UK Biobank whole exome sequencing dataset^24^ (UKB; n = 428,793) and in whole genome sequencing data from the TOPMed sequencing initiatives for the Atherosclerosis Risk in Community (ARIC; n = 10,570) and Cardiovascular Health Study (CHS; n = 2,790) cohorts.^27^ For the Assessment, Serial Evaluation, and Subsequent Sequelae in Acute Kidney Injury (ASSESS-AKI) cohort, CHIP was assessed using a targeted sequencing panel (TWIST Biosciences). Putative somatic variants meeting previously defined criteria for CHIP^9^ were identified using somatic variant caller Mutect2 and filtered using established filtering criteria.^30^ Subgroups of *DNMT3A* CHIP and non-*DNMT3A* CHIP were defined based on the identity of the mutated gene with the largest variant allele fraction (VAF) per person. Additionally, since ASSESS-AKI CHIP status was ascertained by targeted sequencing – a method that that is highly sensitive to small clones compared to whole exome and whole genome-based detection^30^ – a “large CHIP” subgroup was defined as CHIP variants with a VAF ≥ 10%.

### Outcome ascertainment

For the analyses in UKB, ARIC and CHS, incident AKI was ascertained as hospitalizations or deaths with an ICD code for AKI in any position (ICD-9-CM code 584.x or ICD-10-CM code N17.x).^25,26^ In CHS, the following additional ICD-9 codes were included in the AKI definition: 788.9 (uremia), and 586 (renal failure, not otherwise specified), 39.95 (hemodialysis), 54.98 (peritoneal dialysis), V56.8 (peritoneal), and all incident AKI events underwent manual chart review to ensure they met the definition for AKI.^25^ An additional phenotype of severe AKI requiring dialysis (AKI-D) was defined in the UK Biobank as any AKI event with a procedure code for dialysis (OPCS4 X40.x) within 30 days of AKI. Individuals with baseline eGFR < 15 ml/min/1.73 m^2^ (baseline Cr > 3.0 mg/dL in CHS) or documented ESKD, as well as individuals without baseline kidney function measurement were excluded from all analyses. In all three cohorts, the first AKI event was considered as the incident event and no subsequent events were evaluated. Individuals with known AKI prior to enrollment were excluded from the UKB analysis as events are noted only once per person (a limitation of UKB data), whereas previous AKI was not an exclusion criterion in TOPMed cohorts. For the analyses in the ASSESS-AKI cohort, we employed the primary study authors’ definitions when examining the following phenotypes and outcomes: AKI cases and non-AKI controls^28^, non-resolving AKI and resolving AKI cases (resolving AKI defined as a decrease in serum creatinine concentration of 26.5 mmol/L or more or 25% or more from maximum in the first 72 hours after AKI diagnosis; non-resolving AKI defined as any AKI not meeting the definition for resolving AKI)^5^, and primary ASSESS-AKI study outcome (i.e., halving of estimated eGFR or end-stage kidney disease (ESKD)).^5^

### Statistical analysis – prospective cohort studies

For the prospective cohort studies, statistical analyses were performed using R version 4.2.1. Associations of CHIP with incident AKI were assessed using Cox proportional hazards regression while adjusting for age, age^2^, sex, baseline eGFR, baseline smoking status, diabetes, and hypertension, and either 10 principal components of ancestry (UKB) or self-reported race/ethnicity (TOPMed cohorts). In subgroup analyses in the UKB, baseline CKD was defined as baseline eGFR < 60 ml/min/1.73m^2^ or an ICD code for CKD stage 3-5 dated prior to enrollment. Individuals were censored at death, ESKD, or end of study follow-up in the incident AKI analyses, and results were meta-analyzed using a fixed-effect model. For ASSESS-AKI, cross-sectional outcomes were analyzed using Chi-square tests as well as logistic regression analyses adjusted for age, sex, baseline creatinine, AKI stage, smoking status, race, and history of diabetes, hypertension, and cardiovascular disease, and the incident primary outcome was assessed using Cox proportional hazards regression adjusting for the same set of covariates.

### Animals

To determine the effect of hematopoietic *Tet2* deficiency in murine models of AKI, bone marrow transplants were performed as previously reported.^31^ Briefly, recipient mice were lethally irradiated with 9 Gy using a cesium γ source. BM cells were harvested from syngeneic donor femurs and tibias. Recipient mice received a total of 5 × 10^6^ BM cells in 0.2 ml medium through retroorbital injection. Mice received either 100% wild type bone marrow or 80% wild type and 20% bone marrow from mice with hematopoietic stem cell-specific *Tet2* deletion (*Vav1-iCre;Tet2*^*-/-*^). All mice were on the C57/Bl6 background. All donor mice had the CD45.2 isotype and recipient mice had the CD45.1 isotype (Extended Data Figure 1a). C57BL/6 wild-type mice, C57B6/J *Tet2*^f/f^ mice with loxP sites flanking *Tet2* exon 3 (Jackson Laboratories strain number: 017573), and C57B6/J Vav1-iCre mice that enables conditional gene knockout in hematopoietic stem cells (strain number: 008610) were bred at Queen’s University (approved University Animal Care Committee protocol 2021-2128) and provided under material transfer agreement to Dr. Raymond Harris (Vanderbilt University); and C57Bl/6 Cd45.1Pep Boy mice were obtained from Jackson Laboratories (strain number: 002014).

Flow cytometry was used to determine effectiveness of engraftment in the chimeras and assessment of clonal expansion of the hematopoietic cells carrying the *Tet2* deletion. Injury studies were performed when the mutant hematopoietic cells were expanded to ∼60% of total (**Extended Data Figure 3b**).^9,10,32^ Male mice were used for all studies. For ischemia/reperfusion studies, mice were subjected to unilateral kidney vascular clamping for 32 minutes with simultaneous contralateral nephrectomy, as previously described.^33^ For unilateral ureteral obstruction (UUO), the left ureter was ligated as previously described.^34^

### Isolation of kidney macrophages

Kidney single cell suspensions were used to isolate kidney macrophages using anti-mouse F4/80 Microbeads and MACS columns (130-110-443, Milteni Biotec Auburn, CA) following the manufacturer’s protocol.

### Quantitative immunofluorescence/immunohistochemistry staining

Kidney tissue was immersed in fixative containing 3.7% formaldehyde, 10 mM sodium m-periodate, 40 mM phosphate buffer, and 1% acetic acid. The tissue was dehydrated through a graded series of ethanols, embedded in paraffin, sectioned (4 μm), and mounted on glass slides. For immunofluorescence staining, the sections were incubated for two rounds of staining overnight at 4°C. Anti-rabbit or mouse IgG-HRP was used as a secondary antibody. Each round was followed by tyramide signal amplification with the appropriate fluorophore (Alexa Fluor 488 tyramide, Alexa Flour 647 tyramide or Alexa Fluor 555 tyramide, Tyramide SuperBoost Kit with Alexa Fluor Tyramides, Invitrogen) according to the manufacturer’s protocols. DAPI was used as a nuclear stain. Sections were viewed and imaged with a Nikon TE300 fluorescence microscope and spot-cam digital camera (Diagnostic Instruments), followed by quantification using Image J software (NIH, Bethesda, MD).

### Immunoblotting analysis

Whole kidney tissue was homogenized with lysis buffer containing 10 mmol/l Tris–HCl (pH 7.4), 50 mmol/l NaCl, 2 mmol/l EGTA, 2 mmol/l EDTA, 0.5% Nonidet P-40, 0.1% SDS, 100 μmol/l Na3VO4, 100 mmol/l NaF, 0.5% sodium deoxycholate, 10 mmol/l sodium pyrophosphate, 1 mmol/l PMSF, 10 μg/ml aprotinin, and 10 μg/ml leupeptin and centrifuged at 15, 000 x g for 20 min at 4°C. The BCA protein assay kit (Thermo Scientific) was used to measure the protein concentration. Immunoblotting was quantitated with Image J software.

### Antibodies

Antibodies used for immunoblots and/or immunolocalization were: CD68 (Abcam ab125212), NLRP3 (ThermoFisher Cat#PAS079740), IL-1β ThermoFisher Cat #P420B), NGAL (R&D Systems Cat # AF1857), KIM-1 (R&D Systems Cat #AF1817), β-actin (Cell Signaling Cat # 4967), CD45.1 (ThermoFisher 17-0453-82), and CD45.2 (ThermoFisher 14-0454-82).

### Quantitative PCR

Total RNAs from kidneys or cells were isolated using Trizol® reagent (Invitrogen). SuperScript IV First-Strand Synthesis System kit (Invitrogen) was used to synthesize cDNA from equal amounts of total RNA from each sample. Quantitative RT-PCR was performed using TaqMan real-time PCR (7900HT, Applied Biosystems). The Master Mix and all gene probes were also from Applied Biosystems. The probes used in the experiments included mouse *Tnf* (Mm99999068), *Il1b* (Mm00434228), *Il6* (Mm00446190), *Ccl3* (Mm00441258), *Ccl2* (Mm00441242), *Il23a* (Mm00518984), *Col1a1* (Mm00801666), *Col3a1* (Mm01254476), *Col4a1* (Mm01210125), *Acta2* (Mm01546133), *Fn* (Mm01256744), *Tgfb1* (Mm00441726), *Ccn2 (Ctgf*, Mm01192932*), Havcr1* (KIM-1 Mm00506686), *Lcn2L* (NGAL Mm01324470), *Vim* (Mm01333430), *cd68*, (Mm03047343), *il10* (Mm004391), *CD209a* (Mm00460067), *cd163* (Mm00474091), *Mrc1* (CD206 Mm01329362), and *Gapdh* (Mm99999915) was used as a normalizer. Realtime PCR data were analyzed using the 2-ΔΔCT method to determine the fold difference in expression.

**Picro-Sirius red stain** was performed according to the protocol provided by the manufacturer (Sigma, St. Louis, MO, USA).

### Kidney tubular injury score

Periodic acid-Schiff (PAS)–stained slides were used to evaluate tubular injury score. Sections were assessed by counting the percentage of tubules that display cell necrosis, loss of brush border, cast formation, and tubule dilation as follows: 0 = normal; 1 = <10%; 2 = 10 to 25%; 3 = 26 to 50%; 4 = 51 to 75%; 5 = >75%. Five fields from each outer medulla were evaluated and scored in a blinded manner by two observers and the results were averaged.

### Statistical analyses – animal experiments

For animal experiments, statistical analyses were performed with GraphPad Prism 9 (GraphpadSoftware® Inc., La Jolla, CA, US). Data are presented as the mean ± S.E.M. Data were analyzed using 2 tailed Student’s t test or two-way ANOVA followed by Tukey’s or Bonferroni’s post hoc tests. A P value below 0.05 was considered significant. For each set of data, at least 6 different animals were examined for each condition. Collection, analysis, and interpretation of data were conducted by at least 2 independent investigators.

### Study approval

Access to the UK Biobank dataset was provided under application 43397. Local approval for secondary analyses of the data was obtained from the Vanderbilt University Medical Center institutional review board. The Vanderbilt University Medical Centre animal care committee approved all animal study procedures.

## Notes

### Competing Interest Statement

The authors have declared no competing interest.

## References

1. Susantitaphong, P. et al. World Incidence of AKI: A Meta-Analysis. Clinical Journal of the American Society of Nephrology 8, 1482 (2013).

2. Rewa, O. & Bagshaw, S. M. Acute kidney injury—epidemiology, outcomes and economics. Nat Rev Nephrol 10, 193–207 (2014).

3. Lewington, A. J. P., Cerdá, J. & Mehta, R. L. Raising awareness of acute kidney injury: a global perspective of a silent killer. Kidney Int 84, 457–467 (2013).

4. Khwaja, A. KDIGO clinical practice guidelines for acute kidney injury. Nephron Clin Pract 120, c179–184 (2012).

5. Bhatraju, P. K. et al. Association Between Early Recovery of Kidney Function After Acute Kidney Injury and Long-term Clinical Outcomes. JAMA Netw Open 3, e202682–e202682 (2020).

6. Steensma, D. P. et al. Clonal hematopoiesis of indeterminate potential and its distinction from myelodysplastic syndromes. Blood 126, 9–16 (2015).

7. Genovese, G. et al. Clonal hematopoiesis and blood-cancer risk inferred from blood DNA sequence. N. Engl. J. Med. 371, 2477–2487 (2014).

8. Jaiswal, S. et al. Age-related clonal hematopoiesis associated with adverse outcomes. N. Engl. J. Med. 371, 2488–2498 (2014).

9. Jaiswal, S. et al. Clonal Hematopoiesis and Risk of Atherosclerotic Cardiovascular Disease. New England Journal of Medicine 377, 111–121 (2017).

10. Fuster, J. J. et al. Clonal hematopoiesis associated with TET2 deficiency accelerates atherosclerosis development in mice. Science 355, 842–847 (2017).

11. Dorsheimer, L. et al. Association of Mutations Contributing to Clonal Hematopoiesis With Prognosis in Chronic Ischemic Heart Failure. JAMA Cardiol 4, 25–33 (2019).

12. Bhattacharya, R. et al. Clonal Hematopoiesis Is Associated With Higher Risk of Stroke. Stroke 53, 788–797 (2022).

13. Miller, P. G. et al. Association of clonal hematopoiesis with chronic obstructive pulmonary disease. Blood 139, 357–368 (2022).

14. Bolton, K. L. et al. Clonal hematopoiesis is associated with risk of severe Covid-19. Nat Commun 12, 5975 (2021).

15. Wong, W. J. et al. Clonal haematopoiesis and risk of chronic liver disease. Nature 616, 747–754 (2023).

16. Hecker, J. S. et al. CHIP and hips: clonal hematopoiesis is common in patients undergoing hip arthroplasty and is associated with autoimmune disease. Blood 138, 1727–1732 (2021).

17. Agrawal, M. et al. TET2-mutant clonal hematopoiesis and risk of gout. Blood 140, 1094–1103 (2022).

18. Fuster, J. J. et al. TET2-Loss-of-Function-Driven Clonal Hematopoiesis Exacerbates Experimental Insulin Resistance in Aging and Obesity. Cell Reports 33, 108326 (2020).

19. Pasupuleti, S. K. et al. Obesity induced inflammation exacerbates clonal hematopoiesis. J Clin Invest (2023) doi:10.1172/JCI163968.

20. Wang, Y. et al. Tet2-mediated clonal hematopoiesis in nonconditioned mice accelerates age-associated cardiac dysfunction. JCI Insight 5,.

21. Lee, S. et al. Distinct macrophage phenotypes contribute to kidney injury and repair. J Am Soc Nephrol 22, 317–326 (2011).

22. Li, L. et al. The chemokine receptors CCR2 and CX3CR1 mediate monocyte/macrophage trafficking in kidney ischemia-reperfusion injury. Kidney Int 74, 1526–1537 (2008).

23. Conway, B. R. et al. Kidney Single-Cell Atlas Reveals Myeloid Heterogeneity in Progression and Regression of Kidney Disease. JASN 31, 2833–2854 (2020).

24. Vlasschaert, C., Mack, T., Heimlich, J. B., Bick, A. G. & Niroula, A. A practical approach to curate clonal hematopoiesis of indeterminate potential in human genetic datasets. medRxiv (2022).

25. Mittalhenkle, A. et al. Cardiovascular Risk Factors and Incident Acute Renal Failure in Older Adults: The Cardiovascular Health Study. Clin J Am Soc Nephrol 3, 450–456 (2008).

26. Grams, M. E. et al. Performance and Limitations of Administrative Data in the Identification of AKI. CJASN 9, 682–689 (2014).

27. Bick, A. G. et al. Inherited causes of clonal haematopoiesis in 97,691 whole genomes. Nature 1–7 (2020) doi:10.1038/s41586-020-2819-2.

28. Hsu, C. et al. Post–Acute Kidney Injury Proteinuria and Subsequent Kidney Disease Progression: The Assessment, Serial Evaluation, and Subsequent Sequelae in Acute Kidney Injury (ASSESS-AKI) Study. JAMA Internal Medicine 180, 402–410 (2020).

29. Sano, S. et al. Tet2-mediated Clonal Hematopoiesis Accelerates Heart Failure through a Mechanism Involving the IL-1β/NLRP3 Inflammasome. J Am Coll Cardiol 71, 875–886 (2018).

30. Vlasschaert, C. et al. A practical approach to curate clonal hematopoiesis of indeterminate potential in human genetic datasets. Blood blood.2022018825 (2023) doi:10.1182/blood.2022018825.

31. Zhang, M.-Z. et al. Inhibition of cyclooxygenase-2 in hematopoietic cells results in salt-sensitive hypertension. J Clin Invest 125, 4281–4294 (2015).

32. Sano, S. et al. JAK2 V617F -Mediated Clonal Hematopoiesis Accelerates Pathological Remodeling in Murine Heart Failure. JACC Basic Transl Sci 4, 684–697 (2019).

33. Pan, Y. et al. Myeloid cyclooxygenase-2/prostaglandin E2/E-type prostanoid receptor 4 promotes transcription factor MafB-dependent inflammatory resolution in acute kidney injury. Kidney Int 101, 79–91 (2022).

34. Sasaki, K. et al. Deletion of Myeloid Interferon Regulatory Factor 4 (Irf4) in Mouse Model Protects against Kidney Fibrosis after Ischemic Injury by Decreased Macrophage Recruitment and Activation. J Am Soc Nephrol 32, 1037–1052 (2021).

