## Extended Data for "Clonal Hematopoiesis of Indeterminate Potential is Associated with Acute Kidney Injury"

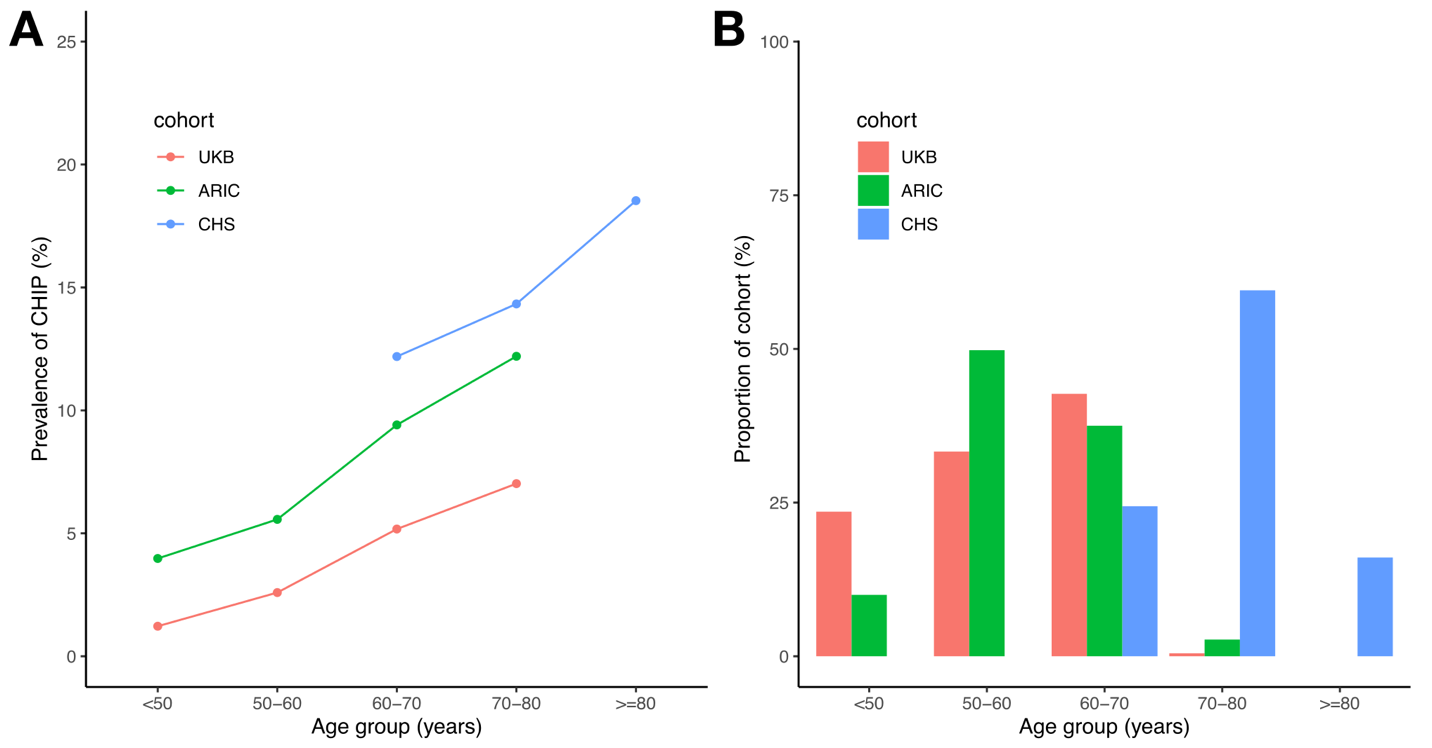

**Extended Data Figure 1.** The distribution of CHIP prevalence by age group across the cohorts included in the study is shown in panel **A**. The proportion of individuals in each age group across the cohorts is shown in panel **B**.

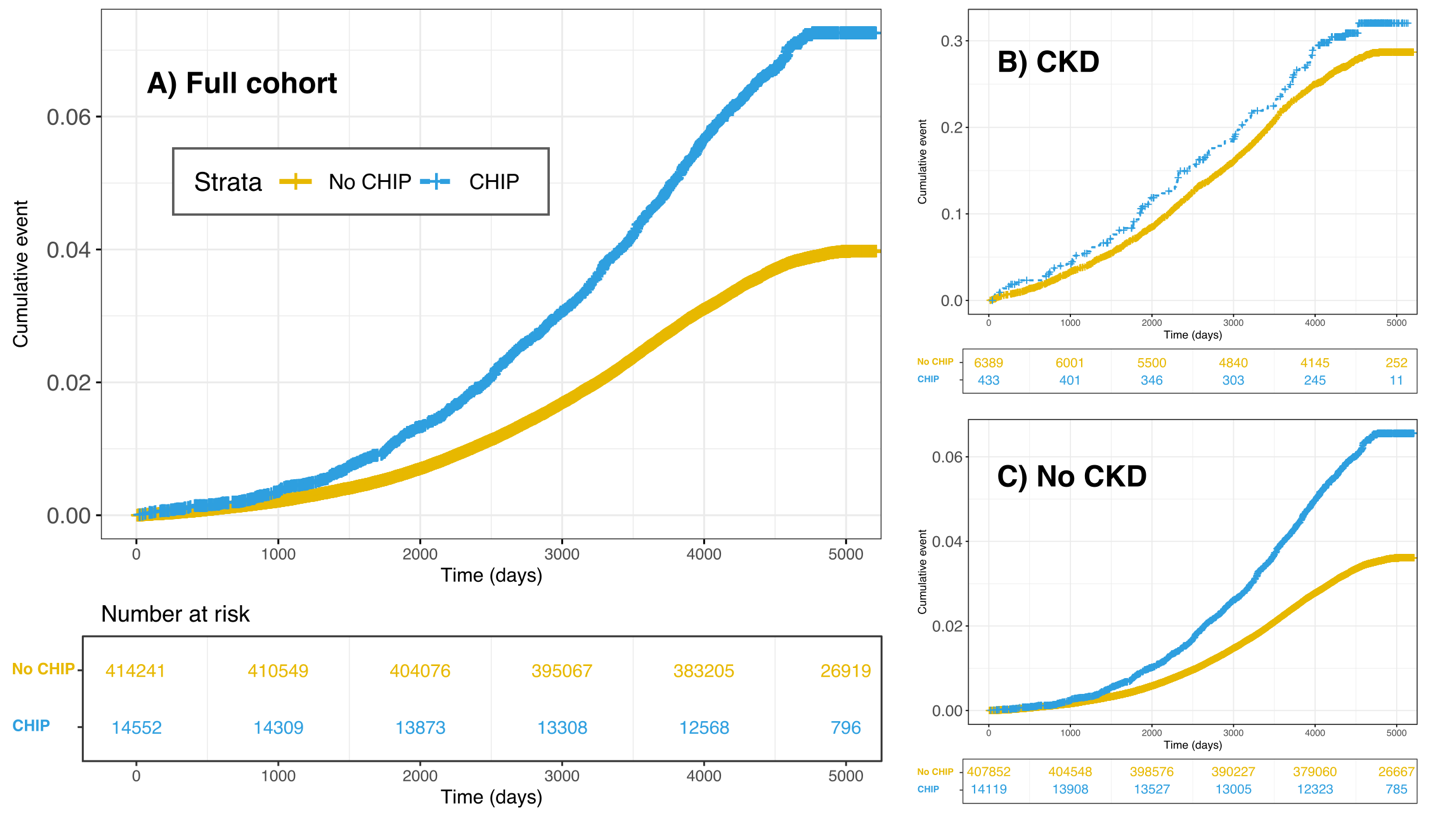

**Extended Data Figure 2.** Kaplan-Meier plots illustrating cumulative AKI events as a function of time in the UKB cohort; shown for (**A**) the whole cohort, (**B**) those with baseline CKD and (**C**) those without baseline CKD. The incidence rate for individuals with CKD and CHIP is 2.88 (events per 100 person-years) and 2.48 for those with CKD without CHIP. For those without CKD, the incidence rate is 0.49 for those with CHIP and 0.27 for those without CHIP.

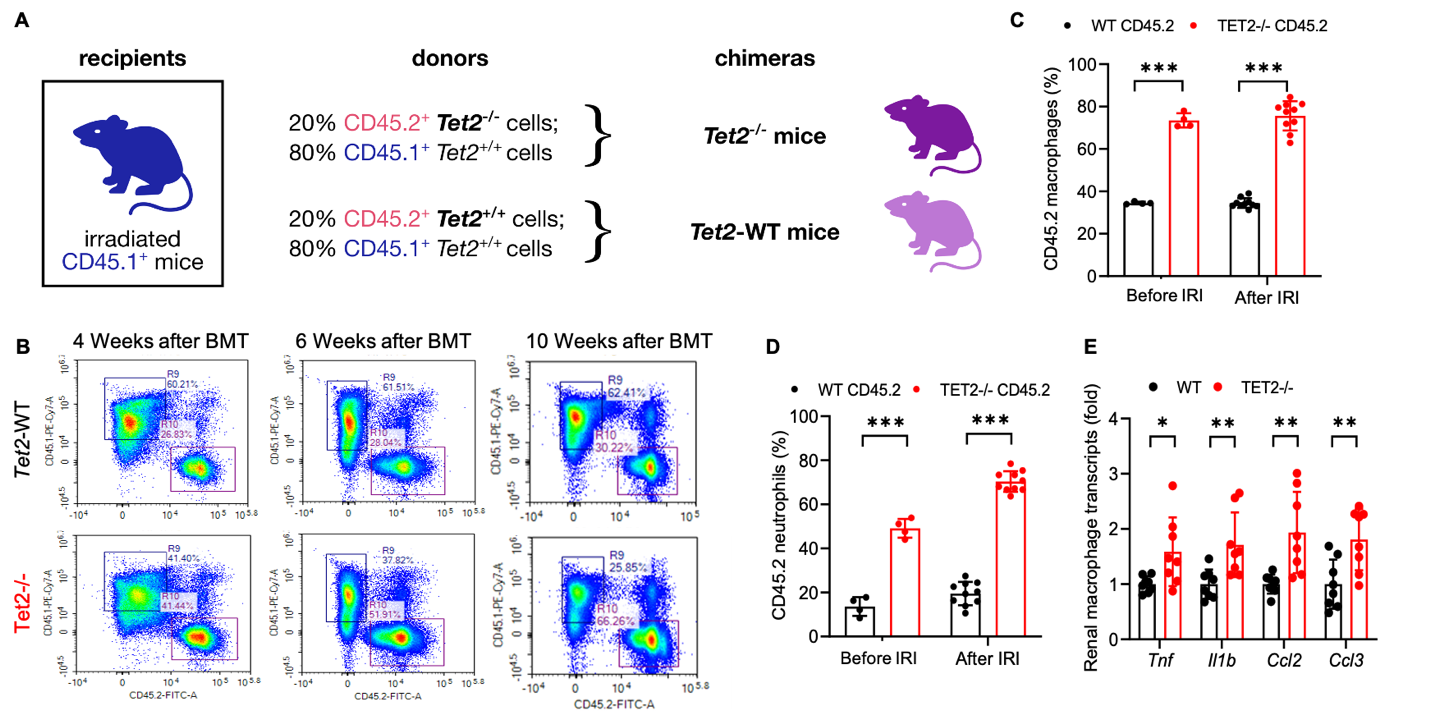

**Extended Data Figure 3** **A**) Schema for production of wild type and *Tet2^-/-^* mice. **B**) Clonal hematopoietic expansion in *Tet2^-/-^* mice. **C&D**) Increased *Tet2^-/-^* macrophages and neutrophils in kidneys without injury and 8 days after ischemic injury. **E**) Increased mRNA of proinflammatory cytokines in kidney macrophages from *Tet2^-/-^* mice without injury. ***p<0.001

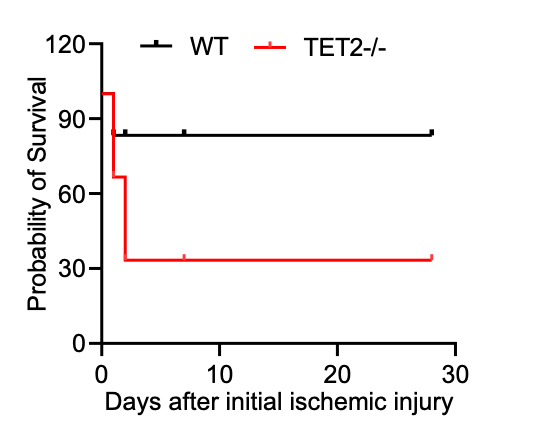

**Extended Data Figure 4.** Survival curve of wild type and *Tet2^-/-^* mice following more severe kidney injury (33.5 minutes of renal pedicle clamping).

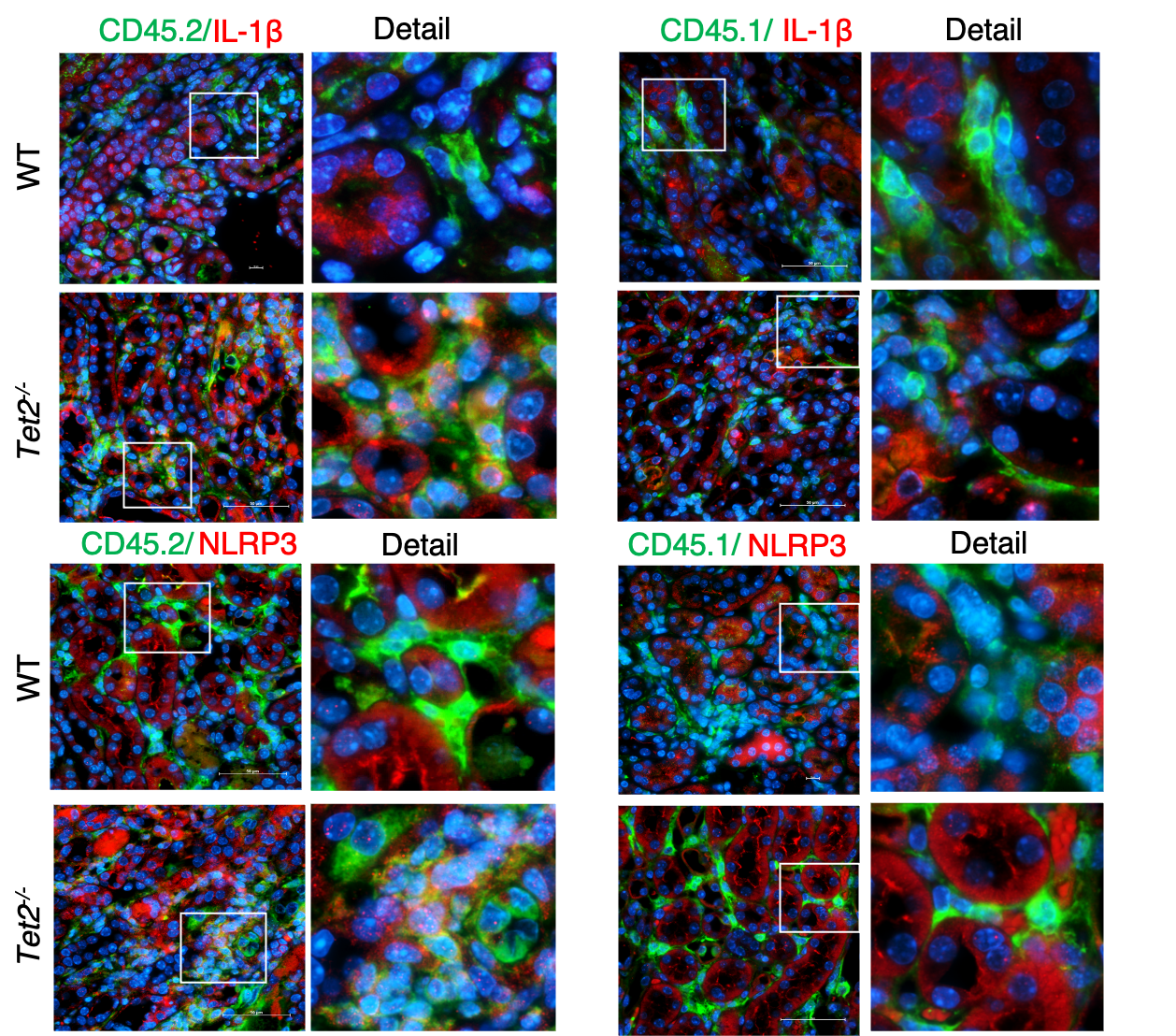

**Extended Data Figure 5.** There was increased colocalization of *Tet2^-/-^* CD45.2 cells but not *Tet2^+/+^* CD45.2 cells nor *Tet2^+/+^* CD45.1 cells with immunoreactive NLRP3 and IL-β.

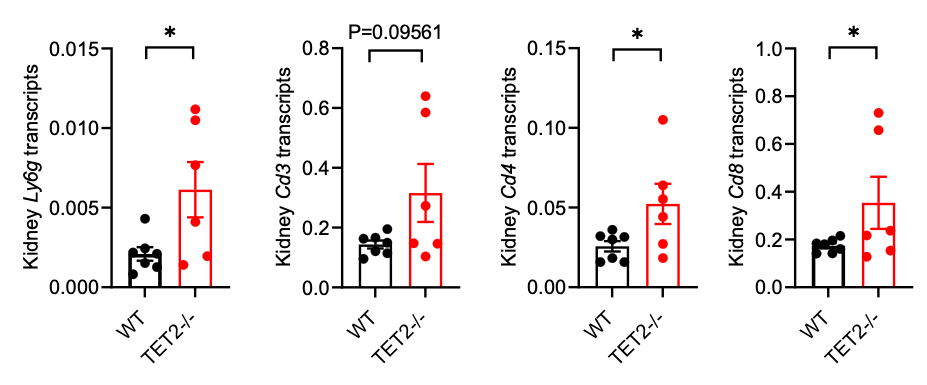

**Extended Data Figure 6.** 28 days after ischemic injury, kidneys of *Tet2^-/-^* mice had increased mRNA markers for neutrophils (*Ly6g*), total T cells (*Cd3*) and CD4 and CD8 T cells. *p<0.05

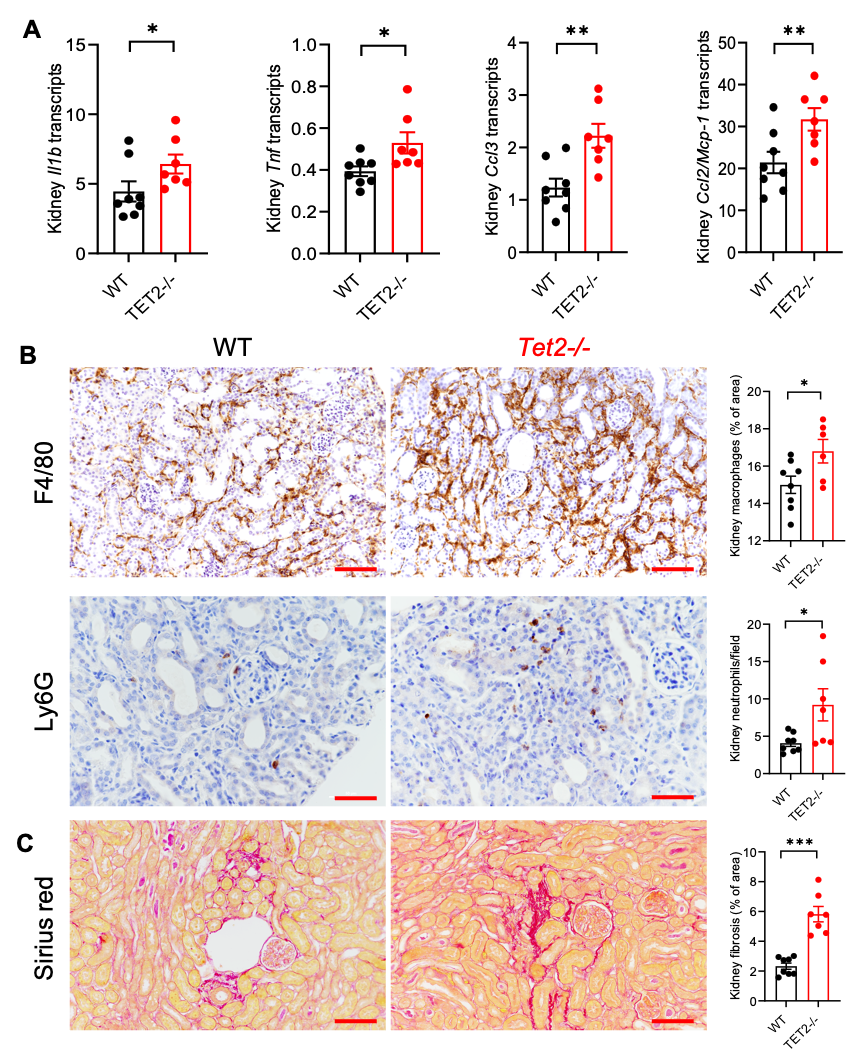

**Extended Data Figure 7.**  7 days after unilateral ureteral obstruction, kidneys of *Tet2^-/-^* mice had **A**) increased mRNA of proinflammatory cytokines, **B**) increased macrophage and neutrophil infiltration and **C**) increased interstitial fibrosis. *p<0.05; **p<0.01; scale bar=50 µm
